# The Effects of Messaging on Expectations and Understanding of Long COVID: An Online Randomised Trial

**DOI:** 10.1101/2022.01.18.22269467

**Authors:** Jaskiran Kaur Bhogal, Freya Mills, Amelia Dennis, Cristina Spoiala, Joanna Milward, Sidra Saeed, Leah Ffion Jones, Dale Weston, Holly Carter

## Abstract

**Objectives:** We examined whether providing different types of information about Long COVID would affect expectations about the illness.

**Design:** A 2 (Illness description: Long COVID vs ongoing COVID-19 recovery) x 2 (Illness uncertainty: uncertainty emphasised vs uncertainty not emphasised) x 2 (Efficacy of support: enhanced support vs basic support) between-subjects randomised online experimental study.

**Setting:** The online platform Prolific, collected in October 2021.

**Participants:** A representative sample of 1110 members of the public in the UK.

**Interventions:** Participants were presented with a scenario describing a positive COVID-19 test result and then presented with one of eight scenarios describing a Long COVID diagnosis.

**Primary and Secondary Outcome Measures:** Various outcome measures relating to illness expectations were captured including: symptom severity, symptom duration, quality of life, personal control, treatment control and illness coherence.

**Results:** We ran a series of 2 × 2 × 2 ANOVAs on the outcome variables. We found a main effect of illness description: individuals reported longer symptom duration and less illness coherence when the illness was described as Long COVID (compared to ongoing COVID-19 recovery). There was a main effect of illness uncertainty: when uncertainty was emphasised, participants reported longer expected symptom duration, less treatment control, and less illness coherence than when uncertainty was not emphasised. There was also a main effect of efficacy of support: participants reported higher personal control and higher treatment control when support was enhanced (compared to basic support). We also found an interaction between illness description and efficacy of support: when support was enhanced, participants reported less illness coherence for Long COVID (compared to ongoing COVID-19 recovery).

**Conclusions:** Communications around Long COVID should not emphasise symptom uncertainty and should provide people with information on how they can facilitate their recovery and where they can access additional support. The findings also suggest that use of the term ongoing COVID-19 recovery, where possible, may reduce negative expectations associated with the illness.

**Strengths and Limitations of this study:**

- This is one of the first experimental designed studies to assess the impact of different types of communication about Long COVID.
- Participants were a UK representative sample, although these findings are not necessarily applicable to all population groups (i.e., ethnic minorities).
- This study is one of the first applications of the IPQ-R in a hypothetical, online experiment, with high reliability.
- This was an online experiment, with hypothetical scenarios and participants with no experience of COVID-19 or Long COVID, therefore outcomes may be different in a real-world context.

## INTRODUCTION

Long COVID, also referred to as Post-COVID-19 syndrome, Post-acute COVID-19, and ongoing symptomatic COVID-19 describes the symptoms of COVID-19 that develop during or after a COVID-19 infection, which continue for more than four weeks and cannot be explained by an alternative diagnosis (1). Common symptoms include fatigue, shortness of breath, muscle ache and difficulty concentrating, with evidence suggesting that such symptoms may be experienced by up to 2.0% of the population (2), affecting day-to-day activities in 64% of those affected (2). Similar post-viral conditions have been observed following other coronavirus infections, such as SARS and MERS, as well as influenza and Ebola (3, 4). Whilst research is ongoing to understand the biological basis for Long COVID, it is well-recognised that psychological factors can also affect physical outcomes associated with a variety of different illnesses (5); it is therefore essential to consider psychological processes alongside biological factors. One such psychological process is one’s expectations of illness progression (6). Illness expectations form part of the Common-Sense Model of Self-Regulation, which posits that illness perceptions influence coping strategies and outcomes (7). These perceptions form one’s illness representation, which are made up of illness label and associated symptoms; illness timeline; physical, cognitive, and social consequences; cause of illness and personal and treatment control. Negative expectations of illness outcomes may make an individual more likely to notice new symptoms, interpret ambiguous sensations unfavourably, and overlook symptom remission (8); they can also influence physiological outcomes (9). Negative expectations that are associated with poorer health outcomes include greater perceived symptom severity (10, 11), greater perceived symptom duration (11), lack of control (including personal control and treatment control) (10-14), and reduced illness coherence (12). It is therefore important to understand factors associated with negative illness expectations, so that these can be reduced.

A key factor that has been shown to increase negative expectations is illness uncertainty. Uncertainty surrounding one’s illness has been linked to increased symptom severity, lack of personal control, decline in mental health and diminished quality of life, amongst other outcomes (10). The Uncertainty in Illness Theory suggests that uncertainty is related to four factors: 1) ambiguity concerning the illness; 2) complexity of treatment, 3) lack of or inconsistent information about diagnosis and severity and 4) unpredictability of prognosis (15). Another factor associated with negative expectations is the way in which an illness is defined (16). Novel illness definitions should be avoided until scientific evidence and medical consensus supports them (17), and existing case definitions should be used in the meantime. Defining a new syndrome is likely to increase strength of illness identity, which has been associated with poorer quality of life (18). An additional factor that has been shown to increase negative expectations is lack of information about the illness (19), and poor signposting to further resources (20). A lack of signposting to further resources can leave patients unable to find reliable information, making it difficult for them to recognise and monitor their own symptoms and access further advice and support (19). Indeed, patients want to be informed of disease modifying therapies, service entitlement and a treatment plan (19), which can promote adherence to suggested treatment and leading to better outcomes (21, 22).

Illness uncertainty, illness definition, and low support can therefore all contribute to negative expectations, which are associated with poorer health outcomes (23). Communication around these three aspects is likely to play a key role in shaping illness expectations associated with Long COVID, and therefore affect health outcomes. Specifically, the extent to which people experience Long COVID symptoms may be affected by 1) the extent to which the likely prognosis is communicated to be uncertain; 2) the definition of Long COVID (i.e., whether the symptoms are described as ongoing COVID-19 recovery or redefined as a new syndrome e.g., Long COVID); 3) the extent to which those experiencing symptoms are given appropriate information and support. To examine the effect of different communication strategies on expectations of Long COVID, the current study used an online experiment to manipulate the information given relating to illness uncertainty (uncertainty emphasised vs uncertainty not emphasised), illness definition (Long COVID vs ongoing COVID-19 recovery), and amount of support provided (enhanced description of support vs basic support). Outcomes included expectations about various different aspects of Long COVID (symptom severity, symptom duration, quality of life, personal control, treatment control, illness coherence), with expectations being more negative to the extent that expected symptom duration and severity were increased, and expected quality of life, personal control, treatment control, and illness coherence were reduced.

### Primary Hypotheses

1. Illness description: We hypothesised that participants would report more negative expectations when symptoms were described as Long COVID rather than ongoing COVID-19 recovery.
2. Illness uncertainty: We hypothesised that participants would report more negative expectations when uncertainty was emphasised compared to when uncertainty was not emphasised.
3. Efficacy of support: We hypothesised that participants would report more negative expectations when basic support was described compared to enhanced support.

### Secondary Hypotheses

We also explored the interaction between illness description, illness uncertainty and efficacy of support.

## METHOD

This project followed the Consolidated Standards of Reporting Trials (CONSORT) reporting guideline (24) (The CONSORT checklist for each item is included in Supplementary File 1).

### Design

This study was an online survey experiment with a between-subjects design with a 2 (Illness description: Long COVID vs ongoing COVID-19 recovery) × 2 (Illness uncertainty: uncertainty emphasised vs uncertainty not emphasised) × 2 (Efficacy of support: enhanced support vs basic support) structure. This resulted in eight conditions, with each participant being randomly allocated to one condition. The main outcome measure was expectations regarding Long COVID, including expectations of symptom duration, symptom severity, quality of life, personal control, treatment control and illness coherence. The study was pre-registered, with the full protocol, at https://www.osf.io/7yq56

### Participants

Participants were invited to complete the survey if they met the inclusion criteria of being older than 18 years old, residing in the UK, not having had a confirmed COVID-19 positive test result, and being fluent in English. We excluded those that had a positive COVID-19 test result to avoid results being influenced by participants’ prior personal experiences of COVID-19 or Long COVID. Power calculations using G*Power 3.1 (25) required a sample size of 1014 for a 2 × 2 × 2 design, achieving 95% power to detect a small effect size (f=0.1). To account for participants failing the attention check question (estimated to be 10%), we aimed to recruit 1120 participants. We recruited 1129 participants, with 19 participants removed due to not completing all measures (*n*=7) or failing the attention check (*n*=12), leaving a final sample size of 1110. We recruited participants through Prolific with monetary compensation of £1.25 based on an expected duration of 10 minutes to complete the survey, according to Prolific’s recommended hourly rate. The survey was piloted with 16 participants to check randomisation procedures, question coherence, obtain any feedback.

### Patient and Public Involvement

We carried out stakeholder involvement in the form of discussions with GPs (*n*=5) to gain an insight into the structure of a typical GP consultation, particularly in relation to cases of suspected Long COVID. We did not carry out patient and public involvement.

### Measures

#### Scenarios

Participants were presented with a hypothetical two-part scenario, developed by the research team based on information available from the NHS (26), National Institute for Health and Care Excellence (1, 27-29), Office for National Statistics (2) and discussions with GPs. The first part of the scenario, which all participants received, described a positive COVID-19 test result. This part of the scenario was designed to set the scene for the second part of the scenario. The second part of the scenario involved a hypothetical GP appointment which contained information relating to: illness description (Long COVID or ongoing COVID-19 recovery); illness uncertainty (emphasised or not emphasised); and available support (enhanced or basic), which was in line with management and referral guidance (29). This resulted in eight variations of the second part of the scenario (see Table 1). Participants were asked to imagine themselves in the given scenarios and answer questions relating to their expectations of the illness, based on the information provided. See Supplementary File 2 for a copy of the scenarios used during the study.

**Table 1.** Overview of Conditions

| Scenario | Condition | *n* | Participants<br>excluded due to<br>attention checks<br>(*n*) | Participants<br>excluded due<br>incompletion<br>(*n*) |
| --- | --- | --- | --- | --- |
| Scenario 1 | Long COVID + Uncertainty<br>Emphasised + Basic Support<br>Condition | 137 | 3 | 1 |
| Scenario 2 | Long COVID + Uncertainty<br>Emphasised + Enhanced<br>Support Condition | 137 | 3 | 1 |
| Scenario 3 | Long COVID + Uncertainty<br>Not Emphasised + Basic<br>Support Condition | 140 | 1 | 0 |
| Scenario 4 | Long COVID + Uncertainty<br>Not Emphasised + Enhanced<br>Support Condition | 140 | 0 | 1 |
| Scenario 5 | Ongoing COVID-19<br>Recovery + Uncertainty<br>Emphasised + Basic Support<br>Condition | 139 | 1 | 1 |
| Scenario 6 | Ongoing COVID-19<br>Recovery + Uncertainty<br>Emphasised + Enhanced<br>Support Condition | 138 | 3 | 1 |
| Scenario 7 | Ongoing COVID-19<br>Recovery + Uncertainty Not<br>Emphasised + Basic Support<br>Condition | 139 | 0 | 2 |
| Scenario 8 | Ongoing COVID-19<br>Recovery + Uncertainty Not<br>Emphasised + Enhanced<br>Support Condition | 140 | 1 | 0 |

#### Primary Outcome Measures

The validated Revised Illness Perception Questionnaire (IPQ-R) (30) was used to measure expected symptom severity, symptom duration, personal control, treatment control and illness coherence. The authors of the IPQ-R encourage that the questions be adapted to suit particular illnesses (30), therefore, the questions were adapted from the original present tense to be in the conditional tense, based on expectations, for example “I would expect my symptoms to come and go in cycles”.

Expected symptom severity: For expected severity, consequences (α= 0.80) and emotional representation (α= 0.85) were measured using the “Consequences” and “Emotional Representation” sub-scales of the IPQ-R. We added two additional questions to the Emotional Representation sub-scale to assess expected severity: “having received this diagnosis, I would expect my symptoms to be serious;” “I would be concerned about my symptoms.”

Expected symptom duration: Expected symptom duration (α=0.86) was measured using the “Timeline Acute/Chronic” sub-scale of the IPQ-R. Participants were also asked an additional question regarding how much longer they would expect to experience their symptoms for with options including less than a month; 1 month; 2-3 months; 4-6 months; 7-9 months; 9-12 months; 12+ months. This question was assessed independently from the scale.

Expected personal control: personal control (α =0.83) assessed participants’ expected control over their symptoms. This was measured using adapted questions from the ‘Personal Control’ sub-scale of the IPQ-R.

Expected treatment control: Treatment control (α= 0.83) assessed participants’ perceptions of efficacy of treatment options. This was measured using adapted questions from the ‘Treatment Control’ sub-scale of the IPQ-R.

Expected illness coherence: Illness coherence (α=0.90) assessed how the illness was understood by participants. This was measured using the ‘Illness Coherence’ sub-scale of the IPQ-R.

Expected quality of life: An adapted version of the World Health Organisation’s Quality of Life questionnaire (31) was used to measure quality of life (α=0.81). Four questions were presented as statements, with participants asked to rate their agreement.

All questions were presented as statements and participants were asked to rate their agreement on a Likert scale from 1 (Strongly Disagree) to 5 (Strongly Agree).

#### Manipulation Checks

The following manipulation checks were asked to ensure that the conditions were perceived as intended. To assess whether participants considered their symptoms to be associated with Long COVID or ongoing COVID-19 recovery, participants were asked: “Considering the information that you have just read, what do you think is the primary cause of your symptoms?” and given the options of 1) ongoing COVID-19 recovery; 2) Long COVID; 3) Another health condition; 4) Don’t know. The first two options for this question were randomised to prevent an order effect. To assess participants’ expectations of symptom uncertainty (α=0.81), an adapted version of the ‘Timeline Cyclical’ theme from the IPQ-R (35) was used, whereby the original present tense was changed to be in the conditional tense, based on expectations. To assess the perceived efficacy of support (α=0.63), participants were asked to rate their agreement with the following statements: “I would find the support provided by the GP helpful” and “I would know what support was available for my diagnosis”. To check that participants could visualise the scenarios (α= 0.76), participants were asked to rate their agreement with the following statements “I was able to imagine this situation well” and “I was able to emotionally engage with this situation.” All questions were presented as statements, in a randomised order, and participants were asked to rate their agreement on a Likert scale from 1 (Strongly Disagree) to 5 (Strongly Agree).

#### Additional Questions

Participants were required to complete an attention check question to check their engagement with the scenarios and the study. Participants were also asked questions relating to their demographic information (age, gender, ethnicity, education, and UK region), their existing health conditions, whether they knew someone with Long COVID and, if so, what their symptoms were and how long they lasted. These variables were collected to understand if they might influence the participants’ responses to questions on negative expectations.

### Procedure

The data was collected between 14^th^ – 16^th^ October 2021. After consenting to the study, participants were presented with the first scenario asking them to imagine that they had received a positive COVID-19 test result. Participants were then asked to answer questions about expected symptom duration and severity. They were then randomly assigned through Qualtrics to receive one of the eight scenarios describing a Long COVID diagnosis. Participants and researchers were blinded to the randomisation. After the scenario, participants completed the measures in the following order: visualisation check, illness coherence, manipulation checks (illness uncertainty and efficacy of support), expected symptom severity (consequences and emotional representation), expected symptom duration, expected personal control, expected treatment control, expected quality of life, manipulation check (illness attribution), attention check, and demographics.

### Analytic Strategy

We first used descriptive statistics to describe demographics and then assessed differences in demographics between conditions using X^2^. Next, we conducted manipulation checks using independent t-tests and X^2^. Finally, we used 2 × 2 × 2 ANOVAs to assess the impact of assigned conditions on symptom severity (consequences and emotional representation), symptom duration, quality of life, personal control, treatment control, and illness coherence.

## RESULTS

### Demographics

A breakdown of the demographic characteristics is presented in Table 2. The majority of participants were white (77.9%) and did not know anyone with Long COVID (63.1%). There were no significant differences in demographics between conditions suggesting that randomisation was successful (see Supplementary File 3).

**Table 2.**
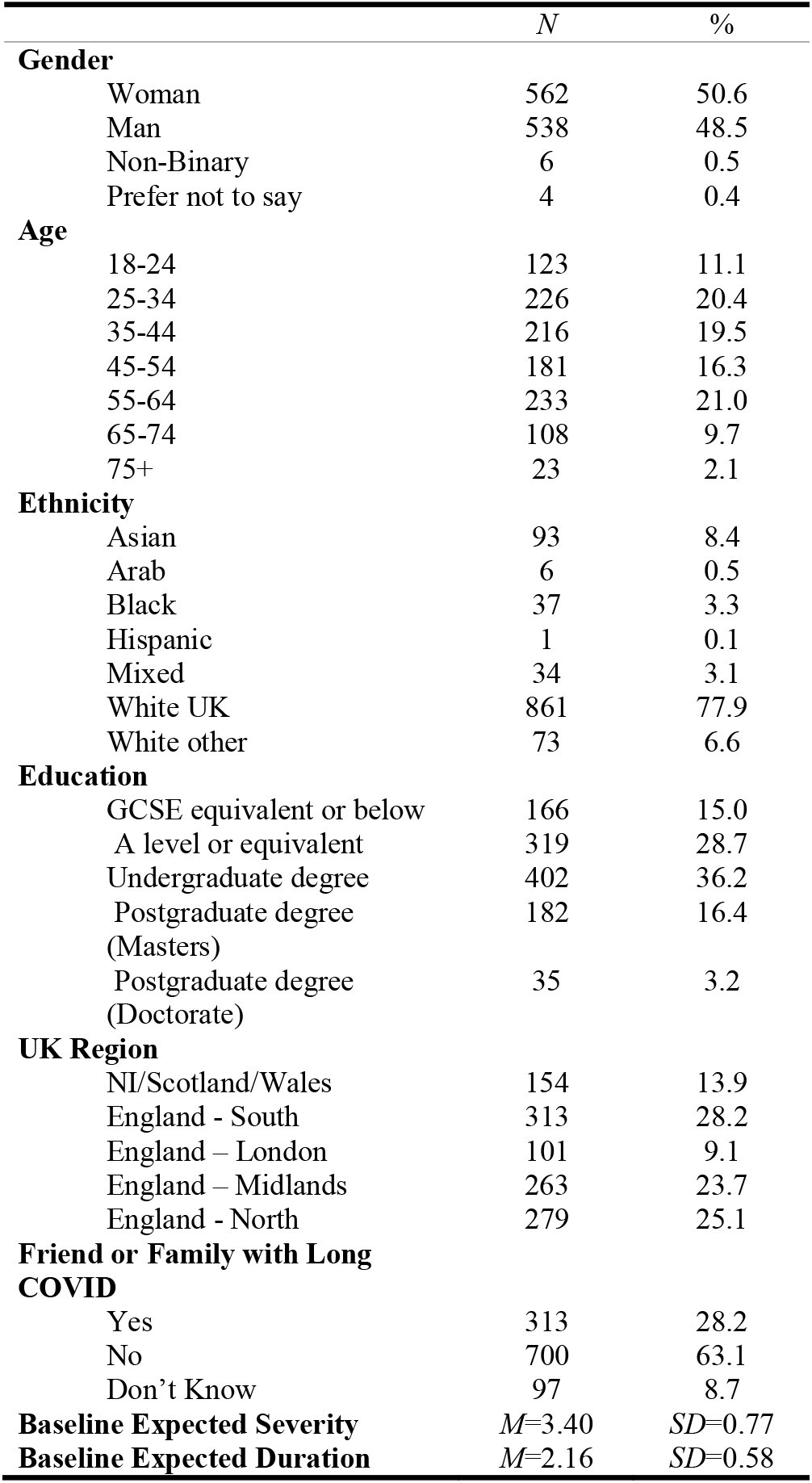
Participant Demographics

### Manipulation Checks

First, we conducted a chi-squared test to examine association between illness description condition (Long COVID vs. ongoing COVID-19 recovery) and attribution of illness (Long COVID vs. ongoing COVID-19 recovery). The relationship was significant, χ2(1) = 60.2, *p* < .001, *V*=0.29, with participants more likely to label the attribution as ongoing COVID-19 recovery in the ongoing COVID-19 recovery conditions (76.3%) compared to the Long COVID conditions (23.7%). Participants were more likely to label the attribution as Long COVID in the Long COVID conditions (58.3%) than in the ongoing COVID-19 recovery conditions (41.7%).

We then ran an independent t-test to assess the effect of the illness uncertainty manipulation (uncertainty emphasised vs. uncertainty not emphasised) on perceived uncertainty. The results showed participants in the uncertainty emphasised conditions (*M*=3.68, *SD*=3.75) reported significantly higher perceived uncertainty than participants in the uncertainty not emphasised conditions (*M*=3.09, *SD*=3.00), *t (*698) =11.7, p<.001, *d*=0.89.

Last, we ran an independent t-test to assess the effect of the efficacy of support manipulation (enhanced support vs. basic support) on perceived efficacy of support. The results showed that participants in the enhanced support conditions (*M*=3.94, *SD*= 4.00) reported significantly greater perceived efficacy of support than participants in the basic support conditions (*M*=3.82, *SD*=4.00), *t (*698) =2.30, *p*=.022, *d*=0.17. Overall, the manipulation checks show that the conditions were effective at inducing illness attribution, perceived uncertainty, and perceived efficacy of support, respectively.

### Effect of Condition

Then we ran a series of 2 (Illness description: Long COVID vs ongoing COVID-19 recovery) × 2 (Illness uncertainty: uncertainty emphasised vs uncertainty not emphasised) × 2 (Efficacy of support: enhanced support vs basic support) ANOVAs to test any differences in expectations of symptom severity (consequences and emotional representation), symptom duration, quality of life, personal control, treatment control, and illness coherence between conditions. The results from the ANOVAs are presented in Table 3 and 4^1^.

**Table 3.** Means and Standard Deviations for Conditions

|  | Long<br>COVID +<br>Uncertainty<br>Emphasised<br>+ Basic<br>Support<br>Condition |  | Long<br>COVID +<br>Uncertainty<br>Emphasised<br>+ Enhanced<br>Support<br>Condition |  | Long<br>COVID +<br>Uncertainty<br>Not<br>Emphasised<br>+ Basic<br>Support<br>Condition |  | Long<br>COVID +<br>Uncertainty<br>Not<br>Emphasised<br>+ Enhanced<br>Support<br>Condition |  | Ongoing<br>COVID-19<br>Recovery +<br>Uncertainty<br>Emphasised<br>+ Basic<br>Support<br>Condition |  | Ongoing<br>COVID-19<br>Recovery +<br>Uncertainty<br>Emphasised<br>+ Enhanced<br>Support<br>Condition |  | Ongoing<br>COVID-19<br>Recovery +<br>Uncertainty<br>Not<br>Emphasised<br>+ Basic<br>Support<br>Condition |  | Ongoing<br>COVID-19<br>Recovery +<br>Uncertainty<br>Not<br>Emphasised<br>+ Enhanced<br>Support<br>Condition |  |
| --- | --- | --- | --- | --- | --- | --- | --- | --- | --- | --- | --- | --- | --- | --- | --- | --- |
|  | M | SD | M | SD | M | SD | M | SD | M | SD | M | SD | M | SD | M | SD |
| Symptom Severity:<br>Consequences | 3.25 | 0.73 | 3.26 | 0.72 | 3.16 | 0.76 | 3.19 | 0.67 | 3.13 | 0.67 | 3.23 | 0.65 | 3.17 | 0.77 | 3.24 | 0.67 |
| Symptom Severity:<br>Emotional<br>Representation | 3.28 | 0.72 | 3.31 | 0.70 | 3.20 | 0.74 | 3.22 | 0.68 | 3.18 | 0.67 | 3.27 | 0.64 | 3.18 | 0.77 | 3.23 | 0.69 |
| Symptom Duration | 3.01 | 0.74 | 3.03 | 0.72 | 2.74 | 0.69 | 2.80 | 0.65 | 2.79 | 0.70 | 2.90 | 0.65 | 2.70 | 0.66 | 2.57 | 0.66 |
| Quality of life | 3.28 | 0.75 | 3.27 | 0.80 | 3.17 | 0.75 | 3.19 | 0.74 | 3.17 | 0.71 | 3.17 | 0.71 | 3.15 | 0.76 | 3.18 | 0.79 |
| Personal Control | 3.15 | 0.73 | 3.33 | 0.65 | 3.14 | 0.71 | 3.24 | 0.68 | 3.07 | 0.70 | 3.20 | 0.65 | 3.10 | 0.70 | 3.18 | 0.73 |
| Treatment Control | 3.14 | 0.77 | 3.22 | 0.69 | 3.20 | 0.74 | 3.27 | 0.72 | 3.10 | 0.72 | 3.20 | 0.73 | 3.24 | 0.70 | 3.35 | 0.73 |
| Illness Coherence | 3.41 | 0.89 | 3.34 | 0.87 | 3.71 | 0.79 | 3.66 | 0.80 | 3.95 | 0.88 | 3.46 | 0.78 | 3.82 | 0.68 | 3.95 | 0.65 |

**Table 4.** ANOVA By Condition on Outcome Variables

|  | Main Effect of Illness Description |  |  | Main effect of Illness Uncertainty |  |  | Main effect of Efficacy of Support |  |  | Interaction Effect of Illness Description and Illness Uncertainty |  |  | Interaction Effect of Illness Description and Efficacy of Support |  |  | Interaction Effect of Illness Uncertainty and Efficacy of Support |  |  | Interaction Effect of Illness Description, Illness Uncertainty, and Efficacy of Support |  |  |
| --- | --- | --- | --- | --- | --- | --- | --- | --- | --- | --- | --- | --- | --- | --- | --- | --- | --- | --- | --- | --- | --- |
| | *F*(1, 692) | *p* | $\eta_p^2$ | *F*(1, 692) | *p* | $\eta_p^2$ | *F*(1, 692) | *p* | $\eta_p^2$ | *F*(1, 692) | *p* | $\eta_p^2$ | *F*(1, 692) | *p* | $\eta_p^2$ | *F*(1, 692) | *p* | $\eta_p^2$ | *F*(1, 692) | *p* | $\eta_p^2$ |
| Symptom Severity: Consequences | 0.26 | .609 | 0.00 | 0.79 | 0.373 | 0.00 | 0.60 | 0.44 | 0.00 | 0.47 | .492 | 0.00 | 1.09 | .296 | 0.00 | 0.26 | 0.607 | 0.00 | 1.11 | .292 | 0.00 |
| Symptom Severity: Emotional Representation | 0.53 | .466 | 0.00 | 2.51 | .113 | 0.00 | 0.36 | 0.547 | 0.00 | 0.13 | .720 | 0.00 | 1.44 | .230 | 0.00 | 0.08 | 0.783 | 0.00 | 1.07 | .300 | 0.00 |
| Symptom Duration | 6.70 | <.001 | 0.01 | 31.78 | <.001 | 0.03 | 0.02 | .656 | 0.00 | 0.26 | .610 | 0.00 | 0.32 | .574 | 0.00 | 1.59 | .208 | 0.00 | 2.90 | .089 | 0.00 |
| Quality of life | 0.58 | .446 | 0.00 | 2.47 | .117 | 0.00 | 0.70 | .402 | 0.00 | 0.19 | .661 | 0.00 | 0.49 | .482 | 0.00 | 0.04 | .843 | 0.00 | 0.29 | .592 | 0.00 |
| Personal Control | 3.29 | .070 | 0.00 | 0.31 | .578 | 0.00 | 8.14 | 0.00 | 0.01 | 0.41 | .523 | 0.00 | 0.14 | .706 | 0.00 | 0.63 | .429 | 0.00 | 0.02 | .878 | 0.00 |
| Treatment Control | 0.13 | .717 | 0.00 | 4.65 | .031 | 0.00 | 4.38 | .037 | 0.00 | 1.07 | .302 | 0.00 | 0.11 | .737 | 0.00 | 0.00 | .987 | 0.00 | 0.03 | .866 | 0.00 |
| Illness Coherence | 4.80 | .029 | 0.00 | 71.44 | <.001 | 0.06 | 0.65 | .420 | 0.00 | 3.47 | .063 | 0.00 | 4.32 | .038 | 0.00 | 0.00 | .987 | 0.00 | 0.02 | .880 | 0.00 |

#### Symptom severity

The results show no main effect of condition or interacting effect of conditions on expected symptom severity in relation to either consequences or emotional representation.

#### Symptom Duration

The results showed two significant main effects on expected symptom duration: illness description and illness uncertainty. In terms of illness description, participants in the Long COVID conditions reported higher expected symptom duration than participants in the ongoing COVID-19 recovery conditions (*p*<.001, *d*=0.23). In regard to illness uncertainty, participants in the uncertainty emphasised conditions reported higher expected illness duration than participants in the uncertainty not emphasised conditions (*p*<.001, *d*=0.34). There was no main effect of efficacy of support or interacting effects.

We also ran three chi squared tests to assess the association between expected symptom duration in months and the three conditions respectively (illness description; illness uncertainty; efficacy of support). There was a significant association between illness uncertainty and months of expected symptom duration, χ2(5) = 46.7, *p* < .001, *V*=0.21. The majority (58.1%) of participants in the uncertainty not emphasised conditions reported expected symptom duration of under 6 months. However, the majority of participants in the uncertainty emphasised conditions (59.3%) expected symptom duration of over 6 months. There was no association between months of expected symptom duration and illness description or efficacy of support.

#### Quality of Life

The results showed no main effect or interacting effects on expected quality of life.

#### Personal Control

The results revealed a main effect of efficacy of support. Participants in the enhanced support conditions reported higher expected personal control than participants in the basic support conditions, *p*=.004, *d*=0.17. No other main effects or interacting effects were significant.

#### Treatment Control

The results showed two significant main effects on expected treatment control: illness uncertainty and efficacy of support. In regard to illness uncertainty, participants in the uncertainty not emphasised conditions reported higher expected treatment control than participants in the uncertainty emphasised conditions (*p*=.031, *d*=0.13). In terms of efficacy of support, participants in the enhanced support conditions reported higher expected treatment control than participants in the basic support conditions (*p*=.037, *d*=0.13). The main effect of illness description was not significant and no significant interacting effects were observed.

#### Illness Coherence

The results showed two significant main effects on illness coherence: illness description and illness uncertainty. In regard to illness description, participants in the ongoing COVID-19 recovery conditions reported significantly higher expected illness coherence than participants in the Long COVID conditions, *p*=.029, *d*=0.13. Additionally, in terms of illness uncertainty, participants in the uncertainty not emphasised conditions reported higher expected illness coherence than participants in the uncertainty emphasised conditions, *p*<.001, *d*=0.51. The interaction between illness description and efficacy of support had a significant effect on expected illness coherence: Participants in the ongoing COVID-19 recovery and enhanced support condition reported higher expected illness coherence than participants in the Long COVID and basic support condition, *p*=.016, *d*=0.26.

### Difference Between Expected Best and Worst Condition

We expected to see a difference between the postulated worst condition (Long COVID + uncertainty emphasised + basic support; Scenario 1) and the postulated best condition (ongoing COVID-19 recovery + uncertainty not emphasised + enhanced support; Scenario 8). We conducted a series of independent t-tests by condition (Scenario 1 vs. Scenario 8) on expected symptom severity (consequences and emotional representation), symptom duration, quality of life, personal control, treatment control, and illness coherence. There were significant differences between Scenario1 and Scenario 8 in expected symptom duration, treatment control, and illness coherence. Participants in Scenario 8 (ongoing COVID-19 recovery + uncertainty not emphasised + enhanced support) reported shorter expected symptom duration, higher expected treatment control, and higher expected illness coherence than participants in Scenario 1. The results from the t-tests are presented in Table 5.

**Table 5.** T-Test Between Best and Worst Scenario

|  | t | p | df | Cohen's<br>d |
| --- | --- | --- | --- | --- |
| Symptom Severity: Consequences | 0.20 | .839 | 275 | 0.02 |
| Symptom Severity: Emotional Representation | 0.61 | .544 | 275 | 0.07 |
| Symptom Duration | 5.20 | <.001 | 275 | 0.63 |
| Quality of life | 0.99 | .324 | 275 | 0.12 |
| Personal Control | -0.30 | .764 | 275 | -0.04 |
| Treatment Control | -2.31 | .021 | 275 | -0.28 |
| Illness Coherence | -5.77 | <.001 | 275 | -0.69 |

## DISCUSSION

We carried out an online experiment to examine the effect of different communication strategies on expectations of Long COVID, including expected symptom duration, expected symptom severity, expected quality of life, expected personal control, expected treatment control, and expected illness coherence. Specifically, we hypothesised that altering the uncertainty of the illness, the name of the condition, and the level of support provided would affect illness expectations, and that expectations would be more negative when: symptoms were described as Long COVID, rather than ongoing COVID-19 recovery; uncertainty of the illness was emphasised; and only basic support was provided.

We found a significant effect of our conditions on four of the six measures of illness expectations (symptom duration, treatment control, personal control, and illness coherence). Participants reported more negative expectations of illness outcomes when the illness was described as Long COVID, when the uncertainty of symptoms was emphasised, and when basic support information was provided rather than enhanced support information. Specifically, when the illness was described as Long COVID rather than ongoing COVID-19 recovery, participants reported longer expected symptom duration and reduced understanding of their illness (illness coherence). When illness uncertainty was emphasised, participants reported longer expected symptom duration, lower expected efficacy of treatment (treatment control), and less understanding of their illness. When information about support was enhanced, compared to basic support information, participants reported higher expected control over their own symptom management (personal control) and higher expected efficacy of treatment. Additionally, participants who received enhanced support information, and an illness description of ongoing COVID-19 recovery, reported greater understanding of their illness than those who received basic support information and an illness description of Long COVID.

Our results broadly supported our hypotheses that emphasising illness uncertainty, defining symptoms as Long COVID, and providing limited information about available support, would increase negative expectations about Long COVID. Thus, we found that changing the way in which Long COVID symptoms and the support available were described affected negative expectations of illness outcomes. Our findings are in line with the extant literature regarding illness uncertainty and health outcomes, as illness uncertainty is associated with increased symptom severity, lack of personal control, decline in mental health, and diminished quality of life, amongst other outcomes in those living with a chronic condition (10). The study also provides support for the Common-Sense Model of Self-Regulation which posits that illness description and symptoms help to shape illness perceptions (7).

However, this study did not find any differences between expected quality of life or expected symptom severity in any of the conditions. One explanation for this could be that the other measures, such as expected symptom duration, illness coherence and personal and treatment control, may be easier to imagine than one’s expected quality of life. This could be due to the hypothetical nature of the experiment, which may have made it difficult for participants to extrapolate to a real-world context. Additionally, quality of life can be influenced by a myriad of factors, including stress or depression (that often accompany physiological conditions (6)), which may also have made it more difficult for participants to accurately imagine their expected quality of life following a Long COVID diagnosis.

### Implications and Recommendations

Our findings suggest that the language used to inform people about Long COVID, including its symptoms and treatment, can play a role in shaping illness expectations. Information which emphasises illness uncertainty, describes symptoms as Long COVID, and fails to provide adequate information about the support available, can increase negative expectations regarding the illness. Given the well-established relationship between negative illness expectations and more adverse health outcomes (6, 12, 23), such negative expectations could have an adverse impact on health outcomes for those experiencing symptoms of Long COVID. The findings from this study therefore emphasise the importance of recognising that the way in which Long COVID is communicated can affect illness expectations (and therefore potentially affect health outcomes) and ensuring that communications around Long COVID are carefully considered. Specifically, communication about Long COVID should provide transparent and factual information about symptoms and available support, whilst not over-emphasising the uncertainty of symptom severity and duration. Whilst it is important not to set up falsely positive expectations, especially given that data are limited, and the illness is relatively new, individuals should be provided with information on how they can personally facilitate their recovery, as well as where they can access additional support. Our findings also suggest that it may be beneficial to use the term ongoing COVID-19 recovery when referring to ongoing COVID-19 symptoms, though this may be challenging given the prominence of the term Long COVID in the narrative around this illness.

### Limitations and Future Research

This study is novel in using an experimental design to assess the effect of different messages about Long COVID on expectations of illness outcomes. A limitation of this research is that the study used a design which required participants to imagine themselves in a hypothetical scenario and report their illness expectations, based on the information provided. While the results are in line with previous research, including that carried out in real-world contexts, caution should therefore be taken when extrapolating current findings to a real-world context. A related limitation is that the hypothetical nature of the experiment meant that we were unable to measure actual health outcomes. While previous research into other conditions demonstrates the relationship between negative illness expectations and adverse health outcomes, the area would benefit from further research to explore the relationship between negative expectations and Long COVID health outcomes, potentially using a longitudinal design. A final limitation is that, although the sample was representative of the UK population, given that the majority UK population are white British, the findings are less generalisable to other ethnicities. This study did not intend to explore any demographic differences, and we found no significant differences in demographic variables between groups, and so it is unlikely that demographic variables affected the outcomes of the study. However, further research is needed to explore the applicability of the current findings to those from other ethnic backgrounds.

## Conclusion

The term Long COVID is used to describe symptoms that develop during or after a COVID-19 infection, which continue for more than four weeks and cannot be explained by an alternative diagnosis. We assessed the impact of different types of information on illness expectations associated with a hypothetical Long COVID diagnosis. We found that describing symptoms as Long COVID, emphasising illness uncertainty, and reducing the description of available support contributed to more negative illness expectations. In light of the well-established link between negative illness expectations and adverse health outcomes, our findings suggest that communications about Long COVID should not emphasise symptom uncertainty and should provide people with information on how they can facilitate their recovery and where they can access additional support. It may also be beneficial to consider using the term ongoing COVID-19 recovery, where possible.

## Supporting information

Protocol

## Data Availability

The data from the current study is available from the corresponding author upon reasonable request.

## Ethics Approval and Consent to Participate

Ethical approval was obtained from Public Health England’s Research and Ethical Governance Group. Before taking part in the study, participants were asked to read an information sheet about the research and complete a consent form to indicate their informed consent to participate. Participants were informed that any information they provided would be confidential, and that they were free to withdraw from the study at any time.

## Transparency Declaration

The lead author (the manuscript’s guarantor) affirms that the manuscript is an honest, accurate, and transparent account of the study being reported; that no important aspects of the study have been omitted; and that any discrepancies from the study as planned (and, if relevant, registered) have been explained.

## Author Contributions

HC conceived the project. JKB, FM, CS, LJ, HC and DW designed and planned the experiment. CS, FM, JKB and SS constructed the experiments on Qualtrics and Prolific. AD conducted statistical analysis. JKB, FM, AD and HC wrote the manuscript with all remaining authors providing comments and feedback. The project was overseen and reviewed by HC and JM.

## Copyright

The Corresponding Author has the right to grant on behalf of all authors and does grant on behalf of all authors, a non-exclusive on a worldwide basis to the BMJ Publishing Group Ltd to permit this article (if accepted) to be published in BMJ editions and any other BMJPGL products and sublicences such use and exploit all subsidiary rights, as set out in our licence.

## Acknowledgments

The authors would like to thank the Behavioural Science and Insights Unit at UKHSA for providing feedback on the questionnaire. In particular, the authors would like to thank Dr Eleonore Batteux for providing support with Qualtrics and Prolific. They would also like to thank the GPs for their time and expertise during the stakeholder engagement.

## Availability of Data

The data from the current study is available from the corresponding author upon reasonable request.

## Footnotes

1 We also ran ANOVAs with gender and age, respectively. We found a main effect of age on illness coherence and an interacting effect of illness description and age on symptom duration and personal control. In terms of gender we found, main effect of gender on emotional representation, symptom duration, personal control, and treatment control and an interacting effect of illness uncertainty conditions, efficacy of support conditions, and gender on symptom duration. See Supplementary Materials for results.

