## Supplementary File 2 for "The Effects of Messaging on Expectations and Understanding of Long COVID: An Online Randomised Trial"

**Supplementary File 1: Scenarios**

***Scenario 1:***

A couple of months after testing positive for COVID-19, you are feeling unwell. You make an appointment to see your GP, where you describe some of the symptoms you are experiencing. You describe symptoms including fatigue (extreme tiredness), shortness of breath, coughing, not being able to think straight (brain fog), chest pain and joint and muscle pain.    

***Scenario 2:***

i. Long COVID + Emphasising Uncertainty + No Enhanced Signposting of Information

Your GP explains that the symptoms you are describing may be due to Long COVID. Your GP explains that at the moment, it is uncertain how long your symptoms will last for or how severe they might be. They also tell you that your symptoms may be unpredictable and that they may affect you in different ways at different times. They explain that your symptoms may get better and then come back and that they may also change over time. They provide you with a leaflet with further information about Long COVID and symptom management.

ii. Long COVID + Emphasising Uncertainty + Enhanced Signposting of Information

Your GP explains that the symptoms you are describing may be due to Long COVID. Your GP explains that at the moment, it is uncertain how long your symptoms will last for or how severe they might be. They also tell you that your symptoms may be unpredictable and that they may affect you in different ways at different times. They explain that your symptoms may get better and then come back and that they may also change over time. They discuss your symptoms with you and provide you with a leaflet containing further information about Long COVID. This includes information on symptoms that you might experience, as well as information about what you can do to manage your symptoms at home. Your GP explains that they will work with you to help you understand and manage symptoms, and that there are support groups and online forums that you can go to for further advice and to help assist your recovery.

iii. Long COVID + Not Emphasising Uncertainty + No Enhanced Signposting of Information

Your GP explains that the symptoms you are describing may be due to Long COVID. They explain that most people will make a full recovery quite quickly, but for some people symptoms can last longer. They provide you with a leaflet with further information about Long COVID and symptom management.

iv. Long COVID + Not Emphasising Uncertainty + Enhanced Signposting of Information

Your GP explains that the symptoms you are describing may be due to Long COVID. They explain that most people will make a full recovery quite quickly, but for some people symptoms can last longer. They discuss your symptoms with you and provide you with a leaflet containing further information about Long COVID. This includes information on symptoms that you might experience, as well as information about what you can do to manage your symptoms at home. Your GP explains that they will work with you to help you understand and manage symptoms, and that there are support groups and online forums that you can go to for further advice and to help assist your recovery.

v. Ongoing COVID-19 Recovery + Emphasising Uncertainty + No Enhanced Signposting of Information

Your GP explains that the symptoms you are describing may be due to ongoing COVID-19 recovery. Your GP explains that at the moment, it is uncertain how long your symptoms will last for or how severe they might be. They also tell you that your symptoms may be unpredictable and that they may affect you in different ways at different times. They explain that your symptoms may get better and then come back and that they may also change over time. They provide you with a leaflet with further information about ongoing COVID-19 recovery and symptom management.

vi. Ongoing COVID-19 Recovery + Emphasising Uncertainty + Enhanced Efficacy of Signposting of Information

Your GP explains that the symptoms you are describing may be due to ongoing COVID-19 recovery. Your GP explains that at the moment, it is uncertain how long your symptoms will last for or how severe they might be. They also tell you that your symptoms may be unpredictable and that they may affect you in different ways at different times. They explain that your symptoms may get better and then come back and that they may also change over time. They discuss your symptoms with you and provide you with a leaflet containing further information about ongoing COVID-19 recovery. This includes information on symptoms that you might experience, as well as information about what you can do to manage your symptoms at home. Your GP explains that they will work with you to help you understand and manage symptoms, and that there are support groups and online forums that you can go to for further advice and to help assist your recovery.

vii. Ongoing COVID-19 Recovery + Not Emphasising Uncertainty + No Enhanced Signposting of Information

Your GP explains that the symptoms you are describing may be due to ongoing COVID-19 recovery. They explain that most people will make a full recovery quite quickly, but for some people symptoms can last longer. They provide you with a leaflet with further information about ongoing COVID-19 recovery and symptom management.

viii. Ongoing COVID-19 Recovery + Not Emphasising Uncertainty + Enhanced Signposting of Information

Your GP explains that the symptoms you are describing may be due to ongoing COVID-19 recovery. They explain that most people will make a full recovery quite quickly, but for some people symptoms can last longer. They discuss your symptoms with you and provide you with a leaflet containing further information about ongoing COVID-19 recovery. This includes information on symptoms that you might experience, as well as information about what you can do to manage your symptoms at home. Your GP explains that they will work with you to help you understand and manage symptoms, and that there are support groups and online forums that you can go to for further advice and to help assist your recovery.
