## Supplementary File 3 for "The Effects of Messaging on Expectations and Understanding of Long COVID: An Online Randomised Trial"

**Supplementary Materials**

**Demographic Differences by Condition**

We ran a series of Chi-Squared association tests and ANOVA by condition on gender, age, ethnicity, education, UK region, friends or family with Long COVID, baseline expected severity and baseline expected duration. The results (see Table 1) showed no significant differences by condition on any demographics.

**The Effect of Demographics**

To assess the influence of age, we ran a series 2 (illness description: Long COVID vs. ongoing COVID-19 recovery) X 2 (uncertainty: uncertainty emphasised vs. uncertainty not emphasised) X 2 (efficacy of support: enhanced support vs. basic support) X 7 (age: 18-24; 25-34; 35-44; 45-54; 55-64; 65-74; 75+) ANOVAs on symptom severity (consequences and emotional representation), symptom duration, quality of life, personal control, treatment control, and illness coherence. All results for these analyses are presented in Table 2. We found a main effect of age on illness coherence. First, illness coherence was lower in 25-34 year olds compared to 55-64 (p<.001, d=-0.50) and 65-74 year olds (-p<.001, d=-0.54). Second, illness coherence was lower in 35-44 year olds compared to 55-64 (p=.003, d=-0.36) and 65-74 year olds (p=.018, d=-0.40). We also found an interaction between illness description and age on symptom duration and personal control. In terms of symptom duration, 18-24 year olds in the Long COVID condition reported higher symptom duration than 25-34 year olds in the ongoing COVID-19 recovery condition (p=.009, d=-0.59) and 35-44 year olds in the ongoing COVID-19 recovery condition (p=.029, d=-0.55). In terms of personal control, we found 75+ year olds in the Long COVID condition reported less personal control than 65-74 year olds in the ongoing COVID-19 recovery condition (p=.018, d=1.30). Although we did find a significant interaction of uncertainty condition and age on quality of life and personal control, post hoc tests for both of these results showed non-significant differences.

To assess the influence of gender, we ran a series 2 (illness description: Long COVID vs. ongoing COVID-19 recovery) X 2 (uncertainty: uncertainty emphasised vs. uncertainty not emphasised) X 2 (efficacy of support: enhanced support vs. basic support) X 2 (gender: woman vs. man) ANOVAs on symptom severity (consequences and emotional representation), symptom duration, quality of life, personal control, treatment control, and illness coherence. We only included women and men as there were not enough non-binary (*n*=6) for their own category. All results for these analyses are presented in Table 3. We found a main effect of gender on emotional representation, symptom duration, personal control, and treatment control. Women reported higher emotional representation (p=.002, d=-0.19), longer symptom duration (p<.001, d=-0.20), but less personal control (p<.001, d=0.23) and less treatment control (p<.001, d=0.21) than men. There was also a significant interaction between illness uncertainty, efficacy of support, and gender on duration. First, in the uncertainty emphasised and enhanced support condition, women reported higher symptom duration than men (p=.002, d=-0.49). Second, men in the uncertainty emphasised and enhanced support condition reported less symptom duration than men (p<.001, d=-0.66) and women (p<.001, d=0.65) in the uncertainty not emphasises and enhances support condition, and men (<.001 d=-0.70) and women (p<.001, d=0.50) in the uncertainty not emphasised and basic support condition.

**Table 1**

*Demographic differences by condition*

|  | Long COVID + Uncertainty Emphasised + Basic Support Condition | | Long COVID + Uncertainty Emphasised + Enhanced Support Condition | | Long COVID + Uncertainty Not Emphasised + Basic Support Condition | | Long COVID + Uncertainty Not Emphasised + Enhanced Support Condition | | Ongoing COVID-19 Recovery + Uncertainty Emphasised + Basic Support Condition | | Ongoing COVID-19 Recovery + Uncertainty Emphasised + Enhanced Support Condition | | Ongoing COVID-19 Recovery + Uncertainty Not Emphasised + Basic Support Condition | | Ongoing COVID-19 Recovery + Uncertainty Not Emphasised + Enhanced Support Condition | | F/X^2^ | p |
| --- | --- | --- | --- | --- | --- | --- | --- | --- | --- | --- | --- | --- | --- | --- | --- | --- | --- | --- |
|  | n | % | n | % | n | % | n | % | n | % | n | % | n | % | n | % |  |  |
| **Gender** |  |  |  |  |  |  |  |  |  |  |  |  |  |  |  |  | 16.4 | .745 |
| Woman | 70 | 51.1 | 61 | 44.5 | 67 | 47.9 | 67 | 47.9 | 61 | 43.9 | 70 | 50.7 | 72 | 51.8 | 70 | 50.0 |  |  |
| Man | 65 | 47.4 | 76 | 55.5 | 71 | 50.7 | 72 | 41.4 | 77 | 55.4 | 66 | 47.8 | 67 | 48.2 | 68 | 48.6 |  |  |
| Non-Binary | 1 | 0.7 | 0 | 0.0 | 0 | 0.0 | 1 | 0.7 | 1 | 0.7 | 1 | 0.7 | 0 | 0.0 | 2 | 1.4 |  |  |
| Prefer not to say | 1 | 0.7 | 0 | 0.0 | 2 | 1.4 | 0 | 0.0 | 0 | 0.0 | 1 | 0.7 | 0 | 0.0 | 0 | 0.0 |  |  |
| **Age** |  |  |  |  |  |  |  |  |  |  |  |  |  |  |  |  | 46.1 | .308 |
| 18-24 | 14 | 10.2 | 17 | 12.4 | 9 | 6.4 | 12 | 8.6 | 18 | 12.9 | 22 | 15.9 | 15 | 10.8 | 16 | 11.4 |  |  |
| 25-34 | 21 | 15.3 | 33 | 24.1 | 30 | 21.4 | 29 | 20.7 | 27 | 19.4 | 27 | 19.6 | 35 | 25.2 | 24 | 17.1 |  |  |
| 35-44 | 25 | 18.2 | 21 | 15.3 | 34 | 24.3 | 29 | 20.7 | 32 | 23.0 | 24 | 17.4 | 25 | 18.0 | 26 | 18.6 |  |  |
| 45-54 | 31 | 22.6 | 16 | 11.7 | 20 | 14.3 | 25 | 17.9 | 25 | 18.0 | 22 | 15.9 | 21 | 15.1 | 21 | 15.0 |  |  |
| 55-64 | 25 | 18.2 | 29 | 21.2 | 31 | 22.1 | 21 | 15.0 | 30 | 21.6 | 29 | 21.0 | 30 | 21.6 | 38 | 27.1 |  |  |
| 65-74 | 16 | 11.7 | 16 | 11.7 | 15 | 10.7 | 19 | 13.6 | 6 | 4.3 | 12 | 8.7 | 10 | 7.2 | 14 | 10.0 |  |  |
| 75+ | 5 | 3.6 | 5 | 3.6 | 1 | 0.7 | 5 | 3.6 | 1 | 0.7 | 2 | 1.4 | 3 | 2.2 | 1 | 0.7 |  |  |
| **Ethnicity** |  |  |  |  |  |  |  |  |  |  |  |  |  |  |  |  | 41.7 | .482 |
| Asian | 12 | 8.8 | 15 | 11.0 | 12 | 8.6 | 8 | 5.7 | 14 | 10.1 | 13 | 9.5 | 7 | 5.1 | 12 | 8.6 |  |  |
| Arab | 0 | 0.0 | 1 | 0.7 | 0 | 0.0 | 1 | 0.7 | 0 | 0.0 | 1 | 0.7 | 1 | 0.7 | 2 | 1.4 |  |  |
| Black | 4 | 2.9 | 4 | 2.9 | 8 | 5.8 | 6 | 4.3 | 3 | 2.2 | 3 | 2.2 | 1 | 0.7 | 8 | 5.7 |  |  |
| Hispanic | 0 | 0.0 | 0 | 0.0 | 0 | 0.0 | 0 | 0.0 | 0 | 0.0 | 0 | 0.0 | 1 | 0.7 | 0 | 0.0 |  |  |
| Mixed | 4 | 2.9 | 3 | 2.2 | 1 | 0.7 | 4 | 2.9 | 6 | 4.3 | 5 | 3.6 | 8 | 5.8 | 3 | 2.1 |  |  |
| White UK | 112 | 82.4 | 106 | 77.9 | 112 | 80.6 | 108 | 77.1 | 103 | 74.1 | 105 | 76.6 | 110 | 79.7 | 105 | 75.0 |  |  |
| White other | 4 | 2.9 | 7 | 5.1 | 6 | 4.3 | 13 | 9.3 | 13 | 9.4 | 10 | 7.3 | 10 | 7.2 | 10 | 7.1 |  |  |
| **Education** |  |  |  |  |  |  |  |  |  |  |  |  |  |  |  |  | 42.6 | .177 |
| GCSE equivalent or below | 27 | 19.7 | 21 | 15.3 | 16 | 11.4 | 24 | 17.1 | 22 | 15.8 | 18 | 13.0 | 24 | 17.3 | 14 | 10.0 |  |  |
| A level or equivalent | 36 | 26.3 | 33 | 24.1 | 54 | 38.6 | 33 | 23.6 | 42 | 30.2 | 42 | 30.4 | 34 | 24.5 | 45 | 36.2 |  |  |
| Undergraduate degree | 48 | 35.9 | 46 | 34.3 | 50 | 35.7 | 52 | 37.1 | 50 | 36.0 | 43 | 31.2 | 53 | 38.1 | 59 | 42.1 |  |  |
| Postgraduate degree (Masters) | 22 | 16.1 | 29 | 21.2 | 17 | 12.1 | 20 | 14.3 | 20 | 14.4 | 30 | 21.7 | 25 | 18.0 | 19 | 13.6 |  |  |
| Postgraduate degree (Doctorate) | 3 | 2.2 | 7 | 5.1 | 2 | 1.4 | 10 | 7.1 | 4 | 2.9 | 3 | 2.2 | 3 | 2.2 | 3 | 2.2 |  |  |
| **UK Region** |  |  |  |  |  |  |  |  |  |  |  |  |  |  |  |  | 23.9 | .684 |
| NI/Scotland/Wales | 14 | 10.2 | 13 | 9.5 | 20 | 14.3 | 30 | 21.4 | 19 | 13.7 | 23 | 16.7 | 17 | 12.2 | 18 | 12.9 |  |  |
| England - South | 44 | 32.1 | 33 | 24.1 | 41 | 29.3 | 37 | 26.4 | 38 | 27.3 | 38 | 27.5 | 43 | 30.9 | 39 | 27.9 |  |  |
| England – London | 12 | 8.8 | 10 | 7.3 | 17 | 12.1 | 11 | 7.9 | 12 | 8.6 | 9 | 6.5 | 13 | 9.4 | 17 | 12.1 |  |  |
| England – Midlands | 33 | 24.1 | 39 | 28.5 | 29 | 20.7 | 35 | 25.0 | 33 | 23.7 | 34 | 24.6 | 28 | 20.1 | 32 | 22.9 |  |  |
| England - North | 33 | 24.1 | 39 | 28.5 | 29 | 20.7 | 35 | 20.0 | 33 | 23.7 | 34 | 24.6 | 28 | 20.1 | 32 | 22.9 |  |  |
| **Friend or Family with Long COVID** |  |  |  |  |  |  |  |  |  |  |  |  |  |  |  |  | 15.0 | .378 |
| Yes | 32 | 23.4 | 36 | 26.3 | 41 | 29.3 | 33 | 23.6 | 45 | 32.4 | 35 | 25.4 | 44 | 31.7 | 47 | 33.6 |  |  |
| No | 93 | 67.9 | 90 | 65.7 | 84 | 90.0 | 94 | 67.1 | 81 | 58.3 | 85 | 61.6 | 88 | 63.3 | 85 | 60.7 |  |  |
| Don’t Know | 12 | 8.8 | 11 | 8.0 | 15 | 10.7 | 13 | 9.3 | 13 | 9.4 | 18 | 13.0 | 7 | 5.0 | 8 | 5.7 |  |  |
| **Baseline Expected Severity** | 3.42 | 0.73 | 3.42 | 0.77 | 3.31 | 0.71 | 3.40 | 0.74 | 3.37 | 0.86 | 3.44 | 0.75 | 3.38 | 0.78 | 3.45 | 0.83 | 0.46 | .867 |
| **Baseline Expected Duration** | 2.18 | 0.64 | 2.18 | 0.60 | 2.16 | 0.54 | 2.17 | 0.57 | 2.10 | 0.60 | 2.16 | 0.52 | 2.19 | 0.58 | 2.15 | 0.56 | 0.33 | .942 |

|  | Main Effect of Age | | | Interaction Effect of Illness Description and Age | | | Interaction Effect of and Illness Uncertainty and Age | | | Interaction Effect of Efficacy of Support and Age | | | Interaction Effect of Illness Description, Illness Uncertainty, and Age | | | Interaction Effect of Illness Description, Efficacy of Support, and Age | | | Interaction Effect of Illness Uncertainty, Efficacy of Support, and Age | | | Interaction Effect of Illness Description, Illness Uncertainty, Efficacy of Support, and Age | | |
| --- | --- | --- | --- | --- | --- | --- | --- | --- | --- | --- | --- | --- | --- | --- | --- | --- | --- | --- | --- | --- | --- | --- | --- | --- |
|  | *F*(1, 692) | *p* | η_p_^2^ | *F*(1, 692) | *p* | η_p_^2^ | *F*(1, 692) | *p* | η_p_^2^ | *F*(1, 692) | *p* | η_p_^2^ | *F*(1, 692) | *p* | η_p_^2^ | *F*(1, 692) | *p* | η_p_^2^ | *F*(1, 692) | *p* | η_p_^2^ | *F*(1, 692) | *p* | η_p_^2^ |
| Symptom Severity: Consequences | 1.27 | .267 | 0.01 | 0.59 | .740 | 0.00 | 2.07 | .054 | 0.01 | 0.74 | .620 | 0.00 | 1.08 | .373 | 0.01 | 0.73 | .622 | 0.00 | 0.23 | .966 | 0.00 | 0.39 | .883 | 0.00 |
| Symptom Severity: Emotional Representation | 0.99 | .431 | 0.01 | 0.77 | .593 | 0.00 | 1.70 | .117 | 0.01 | 0.60 | .727 | 0.00 | 1.22 | .292 | 0.01 | 0.86 | .528 | 0.01 | 0.06 | .999 | 0.00 | 0.45 | 847 | 0.00 |
| Symptom Duration | 0.87 | .520 | 0.01 | 2.24 | .038 | 0.01 | 1.34 | .236 | 0.01 | 0.60 | .734 | 0.00 | 0.34 | .913 | 0.00 | 1.40 | .211 | 0.01 | 1.68 | .122 | 0.01 | 0.58 | .750 | 0.00 |
| Quality of life | 0.59 | .741 | 0.00 | 1.48 | .182 | 0.01 | 2.95 | .007 | 0.01 | 0.42 | .868 | 0.00 | 1.03 | .403 | 0.01 | 0.51 | .803 | 0.00 | 0.24 | .962 | .001 | 0.76 | .601 | .004 |
| Personal Control | 1.68 | .123 | 0.01 | 3.17 | .004 | 0.02 | 2.52 | .020 | .01 | 1.25 | .279 | 0.01 | 1.19 | .308 | 0.01 | 0.90 | .491 | 0.01 | 1.39 | .215 | 0.01 | 1.14 | .336 | 0.01 |
| Treatment Control | 1.90 | .077 | 0.01 | 2.08 | .053 | 0.01 | 1.61 | .140 | 0.01 | 1.10 | .362 | 0.01 | 1.53 | .166 | 0.01 | 0.58 | .747 | 0.00 | 1.61 | .140 | 0.01 | 1.01 | .417 | 0.01 |
| Illness Coherence | 6.72 | <.001 | 0.04 | 0.44 | .852 | 0.00 | 0.49 | .814 | 0.0 | 0.21 | .972 | 0.00 | 1.63 | .136 | 0.01 | 1.58 | .148 | 0.01 | 1.10 | .360 | 0.01 | 1.08 | .369 | 0.01 |

**Table 2**

*The main effect and interacting effects of age*

**Table 3**

|  | Main Effect of Gender | | | Interaction Effect of Illness Description and Gender | | | Interaction Effect of and Illness Uncertainty and Gender | | | Interaction Effect of Efficacy of Support and Gender | | | Interaction Effect of Illness Description, Illness Uncertainty, and Gender | | | Interaction Effect of Illness Description, Efficacy of Support, and Gender | | | Interaction Effect of Illness Uncertainty, Efficacy of Support, and Gender | | | Interaction Effect of Illness Description, Illness Uncertainty, Efficacy of Support, and Gender | | |
| --- | --- | --- | --- | --- | --- | --- | --- | --- | --- | --- | --- | --- | --- | --- | --- | --- | --- | --- | --- | --- | --- | --- | --- | --- |
|  | *F*(1, 692) | *p* | η_p_^2^ | *F*(1, 692) | *p* | η_p_^2^ | *F*(1, 692) | *p* | η_p_^2^ | *F*(1, 692) | *p* | η_p_^2^ | *F*(1, 692) | *p* | η_p_^2^ | *F*(1, 692) | *p* | η_p_^2^ | *F*(1, 692) | *p* | η_p_^2^ | *F*(1, 692) | *p* | η_p_^2^ |
| Symptom Severity: Consequences | 1.95 | .163 | 0.00 | 0.01 | .910 | 0.00 | 1.18 | .277 | 0.00 | 0.07 | .785 | 0.00 | 0.00 | .992 | 0.00 | 0.06 | .802 | 0.00 | 1.64 | .200 | 0.00 | 2.14 | .144 | 0.00 |
| Symptom Severity: Emotional Representation | 9.84 | .002 | 0.01 | 0.14 | .713 | 0.00 | 0.79 | .373 | 0.00 | 0.12 | .729 | 0.00 | 0.01 | .912 | 0.00 | 0.23 | .631 | 0.00 | 0.49 | .485 | 0.00 | 1.53 | .216 | 0.00 |
| Symptom Duration | 11.45 | <.001 | 0.01 | 0.38 | .540 | 0.00 | 2.66 | .103 | 0.00 | 0.57 | .449 | 0.00 | 0.22 | .642 | 0.00 | 0.09 | .764 | 0.00 | 5.33 | .021 | 0.01 | 0.75 | .385 | 0.00 |
| Quality of life | 2.55 | .111 | 0.00 | 0.17 | .677 | 0.00 | 0.47 | .494 | 0.00 | 0.15 | .702 | 0.00 | 0.52 | .470 | 0.00 | 0.00 | .953 | 0.00 | 1.24 | .266 | 0.00 | 1.20 | .274 | 0.00 |
| Personal Control | 14.74 | <.001 | 0.01 | 0.80 | .373 | 0.00 | 1.85 | .175 | 0.00 | 0.02 | .882 | 0.00 | 0.37 | .545 | 0.00 | 1.43 | .231 | 0.00 | 0.18 | .672 | 0.00 | 2.37 | .124 | 0.00 |
| Treatment Control | 12.28 | <.001 | 0.01 | 1.54 | .215 | 0.00 | 0.64 | .422 | 0.00 | 0.18 | .673 | 0.00 | 1.30 | .254 | 0.00 | 3.55 | .060 | 0.00 | 0.19 | .666 | 0.00 | 3.51 | .061 | 0.00 |
| Illness Coherence | 1.19 | .276 | 0.00 | 0.58 | .447 | 0.00 | 2.07 | .150 | 0.00 | 0.72 | .395 | 0.00 | 0.04 | .848 | 0.00 | 2.39 | .122 | 0.00 | 0.13 | .716 | 0.00 | 0.63 | .427 | 0.00 |

*The main effect and interacting effects of gender*
