## Supplementary material for "The Effects of Messaging on Expectations and Understanding of Long COVID: An Online Randomised Trial": Protocol

The effects of messaging on understanding and expectations of post-COVID syndrome: an online experiment

Introduction

Post-COVID syndrome, more commonly referred to as Long COVID, describes the signs and symptoms of COVID-19 that continue or develop weeks or months after the initial illness. The NICE guidelines define post-COVID syndrome as signs and symptoms associated with a COVID-19 infection which continue for more than 12 weeks and cannot be explained by an alternative diagnosis (NICE, 2020). It is estimated that 60,000 people in the UK are currently experiencing symptoms of Long COVID, with the number expected to fluctuate in line with infection rates (NHS England and NHS Improvement Coronavirus 2021). Whilst post-COVID syndrome likely stems from biological processes associated with the initial COVID-19 infection, it has been suggested that psychological factors may also play a role in the progression of the illness; indeed, it is well-recognised that psychological factors can affect physical symptoms associated with a variety of different illnesses, and that psychological approaches can play a key role in effective treatment and management (Pariante, 2021). Whilst research is ongoing to understand the biological basis for post-COVID syndrome, there is also a need to understand relevant psychological processes.

There are various areas of research which may be applicable to an understanding of the psychological factors associated with the development and prognosis of post-COVID syndrome. One relevant body of research explores the ‘nocebo’ effect, whereby people experience negative symptoms following exposure to an inert substance, which can be exaggerated or triggered by psychological mechanisms such as negative expectations (Webster, Weinman & Rubin, 2016; Webster & Rubin, 2019). Such expectations may make an individual more likely to notice new symptoms, interpret ambiguous sensations unfavourably, attribute symptoms to the substance they are taking, and overlook symptom remission (Barsky et al., 2002). Verbal suggestions about symptom likelihood play a key role in the development of negative expectations (Barsky et al., 2002), and increase the likelihood that symptoms will be experienced (Barsky et al., 2002; Meriem et al., 2019). Communicating in such a way that negative expectations are reduced may therefore limit the extent to which people experience adverse symptoms (Meriem et al., 2019).

Expectations of illness progression have also been found to play a role in the development and maintenance of chronic fatigue syndrome and other medically unexplained symptoms, with perceived lack of control, predictability and severity being linked to more adverse outcomes (Clauw et al., 2003; Moss-Morris et al., 1996). These factors are explained within the Uncertainty in Illness theory, which describes uncertainty as being unable to attach meaning to illness-related events and is related to the inability to predict health outcomes (Mishel, 1988). Specifically, uncertainty is related to four factors: 1) ambiguity concerning the state of the illness; 2) complexity of treatment management and healthcare system, 3) lack or inconsistent information about the diagnosis and severity of symptoms and 4) the unpredictability of the disease course and prognosis. This uncertainty has, in turn, been linked to symptom severity, lack of personal control, decline in mental health and diminished quality of life, amongst other outcomes (Wright et al., 2009).

It has been suggested that illness progression could be improved by communicating that symptom are likely to improve or resolve over time (Clauw et al., 2003; Little et al., 2001). It has also been suggested that illnesses should be named based on existing case definitions or overlapping illnesses, rather than with a novel definition that reifies speculative mechanisms before scientific evidence and medical consensus supports them (Clauw et al., 2003). In the case of post-COVID syndrome, it could therefore be beneficial to refer to ongoing COVID-19 recovery, rather than defining a new syndrome (e.g., Long COVID, post-COVID syndrome). Defining a new syndrome in this way is likely to increase the strength of illness identity, another factor that has been associated with poorer quality of life (Moss-Morris, 1996).

In 2004, the UK ‘Better information, better choices, better health’ campaign focused on patients having access to information and health professionals should support their patients in understanding this information (Department of Health, 2006). A lack of signposting further resources can lead patients unable to find reliable information (Swain et al, 2007), with this lack of information making it difficult for patients to recognise and monitor their own symptoms and get further advice and support (Department of Health, 2006). In people with multiple sclerosis, those who reported a lack of information when diagnosed carried out internet searches for more information, while those who reported satisfactory information and treatment were more relaxed about managing their symptoms (Edwards et al., 2008). When receiving a chronic progressive disorder diagnosis, patients want to be informed of disease modifying therapies, service entitlement and a treatment plan (Anestis et al., 2020). Providing patients with these treatment options and clear action plans can increase adherence to suggested treatment and lead to better outcomes (Leventhal et al., 1965; Phillips et al., 2012). Signposting to relevant treatment and support for post-COVID syndrome may therefore reduce patient confusion and enable a plan of action to be developed so that patients are not left feeling confused and so they leave the consultation with a plan of action.

Research into similar syndromes, and an understanding of the mechanisms involved in the nocebo effect, may therefore contribute to an understanding of the development and prognosis of post-COVID syndrome. This enhanced understanding could improve the way in which information about post-COVID syndrome is communicated. Based on research into the nocebo effect and medically unexplained symptoms, there are some key aspects associated with the way in which post-COVID syndrome is communicated that may affect prognosis. Specifically, the extent to which people experience post-COVID syndrome may be affected by 1) the extent to which the likely prognosis is communicated to be uncertain or uncontrollable; 2) the definition of post-COVID syndrome (i.e., whether the symptoms are described as ongoing COVID-19 recovery or redefined as a new syndrome e.g., Long COVID); 3) the extent to which patients are signposted to support. This experiment will therefore examine the effect of different communication strategies on expectations associated with duration, severity, and impact of post-COVID syndrome.

Aims

To investigate the extent to which expectations of symptom duration, symptom severity, and quality of life associated with post-COVID syndrome are affected by the language used to describe post-COVID syndrome, emphasis placed on uncertainty of symptoms, and efficacy of support and information.

Hypothesis

Primary hypotheses

1. Participants are expected to believe that their symptoms will be of shorter duration and/or be less severe and / or they expect a better quality of life if their condition is described as ongoing COVID recovery rather than long COVID.
2. Participants are expected to believe that their symptoms will be of shorter duration and/or be less severe and / or they expect a better quality of life if there is less of an emphasis on uncertainty.
3. Participants are expected to believe that their symptoms will be of shorter duration and / or be less severe and / or they expect a better quality of life if they perceive higher efficacy of support.

Secondary hypotheses

1. Participants are expected to believe that their recovery will be of shorter duration and/or be less severe and expect a better quality of life if there is less of an emphasis on uncertainty **and** if there is an enhanced focus on the efficacy of signposting/information.
2. Participants are expected to believe that their recovery will be of shorter duration and/or be less severe and expect a better quality of life if there is less of an emphasis on uncertainty **and** their condition is described as ongoing COVID recovery rather than long COVID.
3. Participants are expected to believe that their recovery will be of shorter duration and/or be less severe and expect a better quality of life if their condition is described as ongoing COVID recovery rather than long COVID **and** if there is an enhanced focus on the efficacy of signposting/information.
4. Participants are expected to believe that their recovery will be of shorter duration and/or be less severe and expect a better quality of life if their condition is described as ongoing COVID recovery rather than long COVID **and** if there is an enhanced focus on the efficacy of signposting/information **and** if there is less of an emphasis on uncertainty.

Methods

We will follow clinical terminology of ‘post-COVID syndrome,’ however, in the experiment, we will refer to ‘Long COVID’ to keep consistent with the language that participants will be most familiar with.

**Participants**

We will recruit 1120 adults. Gender, age, ethnicity, level of education and UK region will be recorded. We will be screening out participants that are:

- Below the age of 18
- Not living in the UK
- Have had COVID-19 before
- Not fluent in English

Therefore, we are recruiting participants that are:

- Above the age of 18
- Living in the UK
- Have not knowingly had COVID-19
- Are fluent in English

Participants who fail the attention check will be excluded and will not be compensated.

**Power analysis**

A power analysis was conducted for an experiment with a 2 x 2 x 2 design for a between subjects ANOVA to detect a small effect size of 0.1 with 0.95 power. Given that 1014 participants are needed to detect a small effect size, approximately 10% more participants will be recruited to allow for exclusion of any participants who fail the attention check, giving a total of 1120. We used G*Power (version 3.1) to conduct our power analyses.

**Design**

An online experiment with a between-subjects 2 x 2 x 2 design. Participants will read one of eight possible messages about post-COVID syndrome, which will vary:

- how Long COVID is described (Long COVID vs ongoing COVID-19 recovery)
- whether uncertainty of symptoms is emphasised or not (uncertainty vs. no uncertainty)
- efficacy of sources of support/information (enhanced efficacy vs. no enhanced efficacy)

The study is expected to last approximately 10 minutes and will be run on Qualtrics with participant recruitment via Prolific.

**Materials**

All participants will be asked to read a scenario about having received a COVID-19 diagnosis, following this they will answer baseline questions on: expected severity and expected duration. Participants will then be randomised into one of eight groups where they will read one of the eight scenarios. All participants will then answer questions on: understanding of the diagnosis (illness coherence, attribution of symptoms), manipulation checks (perceived efficacy of support, expected uncertainty), expected severity (consequences, general), expected emotional response, expected duration (timeline acute/chronic, timeline cyclical, months), perceived control (personal control, treatment control), expected quality of life, confidence in understanding the questions, demographics, and experience with COVID-19 in the family.

**Procedure**

A representative sample of the UK adult population (based on age, gender, and ethnicity) will be recruited via the online platform Prolific (<https://www.prolific.co/>). Those who have already had confirmed COVID-19 or suspected COVID-19 will be screened out of the experiment.  Before agreeing to take part in the initial survey, participants will view information about the nature of the study. Following this, participants will complete the experiment. On completion, participants will receive a debriefing statement informing them about the nature of the study and signposting them to further resources should they be concerned about any aspect of post-COVID syndrome or the study.

### Outcomes

Primary outcome measures:

- Expectations of symptom severity
- Expectations of symptom duration
- Expectations of quality of life

Secondary outcome measures:

- Perceived control

It is anticipated that outcomes from this experiment will facilitate an understanding of the effect of various messaging on expectations of severity and duration of post-COVID syndrome as well as expectations of quality of life. This will provide valuable insight into how such outcomes will be affected by different messaging around post-COVID syndrome and how messaging should be optimised in the future in order to communicate post-COVID syndrome in the way that causes least harm.

**Funding**

The study will be funded by the Behavioural Science and Evaluation within Health Protection Research Unit. Based on Prolific’s recommendations and comparable experiments (e.g., those conducted by UCL’s Psychology Department), participants will be paid at a rate of £7.50/hour. For a 10-minute experiment, this equates to £1.25. For a total of 1432 participants, including 33% service fees and 20% VAT on these fees, the total cost should be approximately £3,581.18 (£1,790 of which is participant costs).

It will be necessary to pilot the study initially with 16 participants (2 per condition) to ensure the duration estimate is accurate, to ensure it runs correctly and that participants do not experience any problems completing the experiment. It is anticipated that this may incur additional costs if the experiment needs to be adapted after the pilot.

Analysis

To test this, we will conduct a 2 x 2 x 2 between subjects ANOVA to understand the effect of the type of diagnosis that people read on their perception of duration, severity and quality of life. The data will be analysed with SPSS and R.

Project Timeline

The experiment is expected to go live during the second week of July; however, the research team acknowledges that the study may be delayed if adjustments need to be made.

Ethical considerations

The study will ask participants about their expectations and understanding of Long COVID in a hypothetical scenario. Informed consent will be obtained from all participants before they participate in the study. The study will be submitted and reviewed by the PHE Research Ethics and Governance Group. Data will be stored in line with GDPR requirements, and no identifiable information will be recorded. The study will be preregistered on the Open Science Framework.

Data handling

Survey responses will remain anonymous, will be stored on secure PHE servers, and will not be shared outside of the working group, in line with GDPR regulations.
